# EPIC4ND: European Prospective Investigation into Cancer and Nutrition follow-up for neurodegenerative diseases

**DOI:** 10.1101/2025.01.29.25321340

**Authors:** Christina M. Lill, Jan Homann, Olena Ohlei, Karl Smith-Byrne, Vivian Viallon, Jose Maria Huerta, Fanny Artaud, Yujia Zhao, Abigail Britten, Laura Deecke, Valerija Dobricic, Olatz Mokoroa, Marcela Guevara, Dafina Petrova, Miles Trupp, Jara Sabin, Mareike Gröninger, Sandra M. Colorado-Yohar, Sonia Alonso-Martin, Natalia Cabrera-Castro, Sergiu Groppa, Oliver Robinson, Johnni Hansen, Sture Eriksson, Sabina Sieri, Ana Jimenez, Tammy Y. N. Tong, Rudolph Kaaks, Gianluca Severi, Ruth Travis, Nick Wareham, Andréa L. Benedet, Henrik Zetterberg, Andre Franke, Alexis Elbaz, Lars Bertram, Roel Vermeulen, Lefkos Middleton, Giovanna Masala, Carlotta Sacerdote, Susan Peters, Verena Katzke, Pietro Ferrari, Marc J. Gunter, Elio Riboli

**Affiliations:** Institute of Epidemiology and Social Medicine, University of Münster, Münster, Germany; Ageing and Epidemiology Unit (AGE), School of Public Health, Imperial College London, London, UK; Cancer Epidemiology Unit, Nuffield Department of Population Health, University of Oxford, Oxford, UK; International Agency for Research on Cancer (IARC/WHO), Nutrition and Metabolism Branch, 69366, Lyon, France; Department of Epidemiology, Murcia Regional Health Council-IMIB, Murcia, Spain; CIBER Epidemiología y Salud Pública (CIBERESP), Madrid, Spain; Université Paris-Saclay, UVSQ, Inserm, Gustave Roussy, CESP, Villejuif, France; Institute for Risk Assessment Sciences, Utrecht University, Utrecht, The Netherlands; Medical Research Council Epidemiology Unit, University of Cambridge, Cambridge, UK; Lübeck Interdisciplinary Platform for Genome Analytics (LIGA), University of Lübeck, Lübeck, Germany; Epidemiology and Public Health Area, Biodonostia Health Research Institute, San Sebastián, Spain; Sub-Directorate for Public Health and Addictions of Gipuzkoa, Ministry of Health of the Basque Government, San Sebastián, Spain; Instituto de Salud Pública y Laboral de Navarra, Pamplona, Spain; Centro de Investigación Biomédica en Red de Epidemiología y Salud Pública (CIBERESP), Madrid, Spain; Navarra Institute for Health Research (IdiSNA), Pamplona, Spain; Escuela Andaluza de Salud Pública (EASP), Granada, Spain; Instituto de Investigación Biosanitaria ibs.GRANADA, Granada, Spain; Department of Clinical Sciences, Neurosciences, Umeå University, Umeå, Sweden; Division of Cancer Epidemiology, German Cancer Research Center, Heidelberg, Germany; 1st Medical Clinic, University Hospital Mannheim, Mannheim, Germany; 9 enero Research Group on Demography and Health, National Faculty of Public Health, University of Antioquia, Medellin, Colombia; Stem Cells and Aging Group Bioengineering Area, BioGipuzkoa (BioDonostia) Health Research Institute, San Sebastián, Spain; Department of Neurology, Saarland University Clinic, Saarland, Germany; Department of Epidemiology and Biostatistics, School of Public Health, Imperial College London, London, UK; Danish Cancer Institute, Danish Cancer Society, Copenhagen, Denmark; Department of Public Health and Clinical Medicine, Umeå University, Umeå, Sweden; Epidemiology and Prevention Unit, Fondazione IRCCS Istituto Nazionale dei Tumori di Milano, Milan, Italy; Sub Directorate for Public Health and Addictions of Gipuzkoa, Ministry of Health of the Basque Government, San Sebastian, Spain; Epidemiology of Chronic and Communicable Diseases Group, BioGipuzkoa (BioDonostia) Health Research Institute, San Sebastián, Spain; Department of Statistics, Computer Science, Applications “G. Parenti”, University of Florence, Italy; Department of Psychiatry and Neurochemistry, University of Gothenburg, Molndal, Sweden; Clinical Neurochemistry Laboratory, Sahlgrenska University Hospital, Mölndal, Sweden; Department of Neurodegenerative Disease, UCL Institute of Neurology, Queen Square, London, UK; UK Dementia Research Institute at UCL, London, UK; Hong Kong Center for Neurodegenerative Diseases, Clear Water Bay, Hong Kong, China; Wisconsin Alzheimer’s Disease Research Center, University of Wisconsin School of Medicine and Public Health, University of Wisconsin-Madison, Madison, WI, USA; Institute of Clinical Molecular Biology, Christian-Albrechts-University of Kiel, Kiel, Germany; University Medical Centre Utrecht, Utrecht, The Netherlands; Clinical Epidemiology Unit, Institute for Cancer Research, Prevention and Clinical Network (ISPRO), Florence, Italy; Department of Health Sciences, University of Eastern Piedmont, Novara, Italy

## Abstract

The ‘European Prospective Investigation into Cancer and Nutrition’ cohort (EPIC-Europe) is a prospective study including ∼520,000 participants recruited across Europe (1991-2000) with in-depth baseline data on nutritional, lifestyle, medical, and anthropometric variables, and baseline blood samples. Here we introduce EPIC4ND, a case-cohort study within EPIC designed to identify biomarkers predicting a future onset of dementia, Alzheimer’s disease (AD), Parkinson’s disease (PD), and amyotrophic lateral sclerosis (ALS).

EPIC4ND comprises 6,346 initially non-diseased participants (aged 35-80 years, mean age at baseline: 54±9, 65% women) with up to 30 years of follow-up and data on at least one *omics* domain available from pre-disease blood samples. EPIC4ND includes 4,604 subcohort members (4,447 non-cases and 157 incident cases) and 1,742 additional incident cases ascertained from the broader EPIC cohort. Among the incident cases, there are 1,190 dementia cases (818 AD), 534 PD cases, and 199 ALS cases. Additionally, 72 prevalent PD cases and 118 incident Parkinsonism cases are available for comparison. Molecular data generated encompass proteomics, genome-wide DNA methylation, and SNP genotyping with 4,065 EPIC4ND participants (2,497 non-cases, 1,568 incident cases) having data on all three domains. Smaller studies include data on metals, metabolites, and environmental chemicals, while ongoing efforts focus on ultrasensitive targeted biomarker measurements and small RNA sequencing.

Genome-wide association studies and analyses of epidemiological risk factors validate the dataset by confirming many known risk factors. Leveraging these extensive pre-disease multi-layered *omics* data offers a unique opportunity to identify biomarker signatures predicting neurodegenerative diseases and to explore their interplay with epidemiological risk factors.

## The EPIC-Europe cohort

The ‘European Prospective Investigation into Cancer and Nutrition’ cohort (EPIC-Europe) is a multi-centre prospective study including 519,978 participants (70.5% women, mostly aged 35-70 years at baseline) recruited from 23 centers across ten European countries.^1,2^ With approximately 30 years of follow-up, EPIC was originally designed to explore the relationships between diet and cancer, while also having the potential to investigate other chronic diseases. Enrollment at each center occurred between 1991 and 2000. Baseline data were collected on diet, physical activity, reproductive history, alcohol and tobacco use, previous and current illnesses, as well as anthropometric measurements, all following standardized protocols. Blood samples were collected and separated into citrate plasma, serum, buffy coats, and erythrocytes using identical protocols and equipment, and then aliquoted for long-term storage in liquid nitrogen (-196°C) at a central biobank located at the International Agency for Research on Cancer (IARC)/World Health Organization in Lyon, France.^1,2^ Since baseline recruitment, participants have been followed up regularly by data linkage and/or questionnaires at each center. Since the initial enrollment, several working groups have conducted case ascertainments and research projects to investigate the role of lifestyle and dietary factors on a range of outcomes beyond cancer, such as diabetes^3^, cardiovascular disease^4^, inflammatory bowel disease^5^, and neurodegenerative diseases^6–8^. This also offers the possibility to investigate multimorbidity as well as the intersections of risk factors and pathways across chronic diseases.

### Rationale for the new focus and new data collection

Neurodegenerative diseases (ND) such as Alzheimer’s disease (AD), Parkinson’s disease (PD), and amyotrophic lateral sclerosis (ALS) are becoming increasingly prevalent, particularly in aging, industrialized populations, leading to increasing costs for the public health systems. For example, the number of people diagnosed with dementia is projected to rise from 55 million in 2019 to 139 million by 2050 globally. As a result, the costs associated with dementia are expected to drastically increase, too, from 1.3 trillion US dollars annually in 2019 to 2.8 trillion US dollars by 2030 (https://www.alzint.org/u/World-Alzheimer-Report-2023.pdf). Given these trends, the ultimate goal of translational research in the ND field is to develop strategies for preventing these devastating and currently incurable conditions. It is widely recognized that identifying individuals at risk in pre-disease or early disease stages is crucial for improving the success of clinical treatment trials (e.g., ref. ^9^). However, pathophysiological mechanisms commence up to 20 years or more prior to a clinical diagnosis for AD and PD (while the prodromal period for ALS is largely unknown). The current lack of clinically relevant predictive biomarkers makes the identification of high-risk individuals difficult to impossible. To address this, we established the EPIC4ND case-cohort with the aim to develop prediction models for dementia (particularly AD), PD and ALS. This research will be based on multidimensional molecular biomarkers derived from blood samples taken at baseline, i.e., at a pre-disease stage.

### New areas of research

To identify molecular signatures and biomarkers predictive of a future onset of AD/dementia, PD, and ALS, several layers of molecular *omics* data were recently generated in baseline blood samples of up to 6,346 EPIC participants using high-throughput methodologies (**Figure 1**). This includes proteomic data (SomaScan) generated from plasma samples and genomic and epigenomic (i.e., methylomic) data from blood-derived DNA samples (see below). In addition, the generation of transcriptomic (small RNAs) data and the targeted protein biomarker data for amyloid and tau pathology and neurodegeneration (ultrasensitive immunoassays) is currently underway. The new areas of research will focus on: i) identifying novel individual molecular biomarkers for a future onset of AD, PD, and ALS, ii) discovering molecular biomarker signatures by combining multiple markers within and across molecular domains, iii) investigating biomarkers and pathways shared across the investigated ND and in comparison to other chronic diseases, and iv) integrating these signatures with established demographic and lifestyle risk factors for predictive modelling. The EPIC4ND case-cohort will serve as a valuable resource for addressing a range of additional related questions by exploiting and integrating the available high-dimensional data in the framework of the EPIC study: This includes assessing interactions among markers within the same and across different molecular domains, as well as their interplay with demographic, medical, and lifestyle factors. It also involves exploring the role of biological aging through the construction of molecular clocks and uncovering pathophysiological mechanisms underlying our biomarker observations. Additionally, this multi-*omics* resource can be utilized for research on other quantitative phenotypes measured within the corresponding subcohort in EPIC.

**Figure 1.**
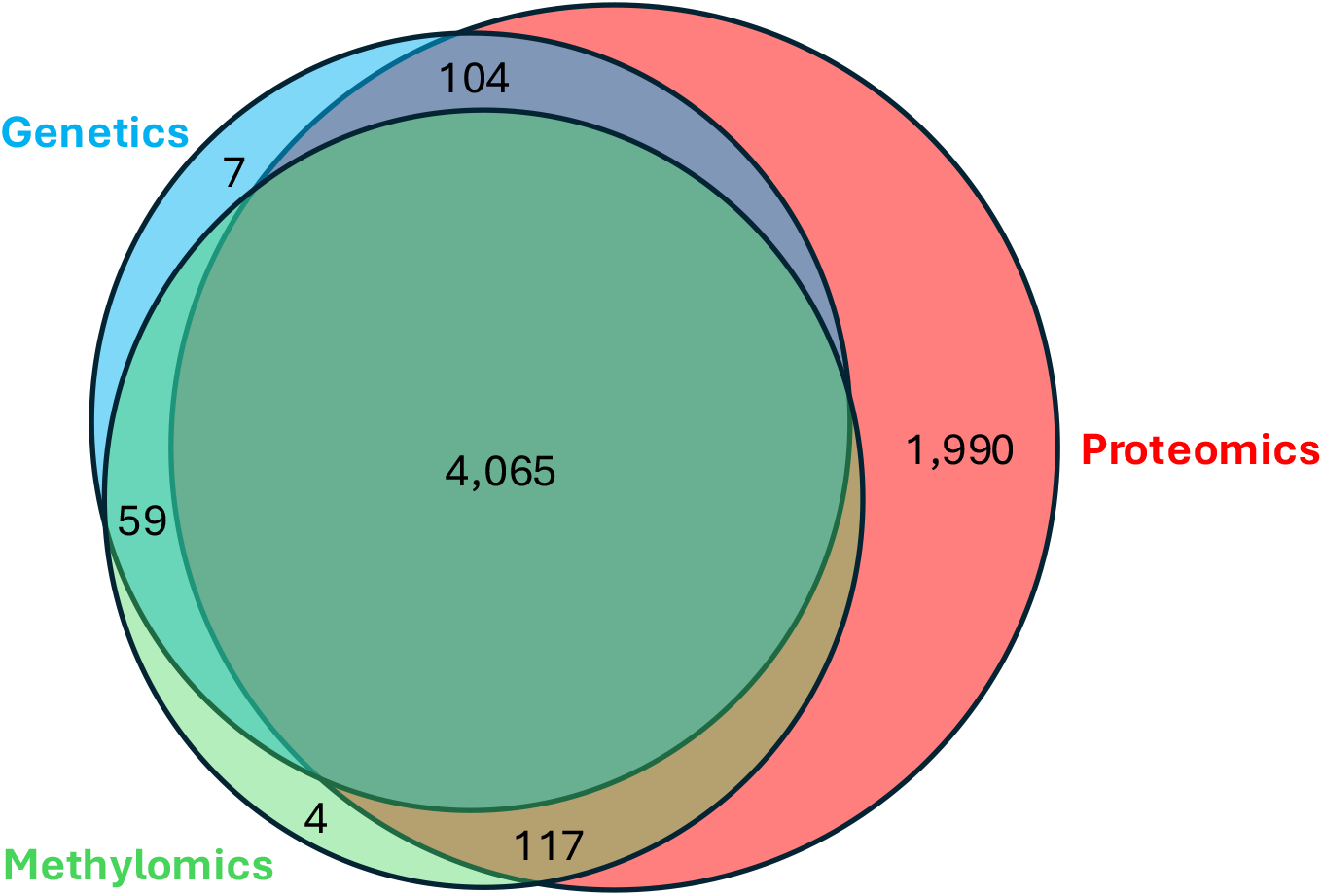
Venn diagram illustrating the omics domains analyzed in the EPIC4ND case-cohort. Legend. This figure depicts the number of samples from the EPIC4ND case-cohort with measurements available for at least one *omics* domain after quality control. Notably, small RNASeq data for 3,992 samples and data on established neurodegenerative disease biomarkers for 1,700 samples of the core EPIC4ND dataset (i.e., those with data from all three *omics* domains) are currently being generated.

### Description of the case-cohort

The EPIC4ND case-cohort comprises a total of 6,346 participants aged 35-80 years at baseline who were recruited as part of the EPIC study in twelve centers across five countries. Quality-controlled data from at least one molecular domain generated from baseline blood samples is available for each EPIC4ND participant. According to the case-cohort design, the 6,346 participants comprised 4,604 subcohort members (thereof 4,447 non-cases from the subcohort that did not have incident dementia, PD, and ALS, as well as 157 incident cases) and 1,742 additional incident cases ascertained from the broader EPIC cohort. Subcohort members were randomly drawn among all participants 35-80 years at baseline from within the existing EPIC-InterAct subcohort^3^. Varying numbers of subcohort members are available for the analyses per disease entity depending on the number of centers contributing cases (**Table 1, Supplementary Tables 1-4**). These centers include Murcia, Andalucía/Granada, Gipuzkoa/San Sebastian, Navarra (all Spain), Turin, Florence, Varese (Italy), Cambridge, Oxford (United Kingdom), Utrecht, Bilthoven (Netherlands), and Heidelberg (Germany). Specifically, dementia cases (n=1,190, thereof 818 AD) were ascertained across all EPIC participants in the four Spanish centers, PD cases (n=534) across all EPIC participants in ten centers (all except Andalucía/Granada and Oxford) across all five countries, and ALS cases (n=199) in all twelve centers (**Table 1, Supplementary Tables 1-4**). Notably, 193 eligible incident ND cases who did not have biomaterial available were excluded from the EPIC4ND case-cohort.

**Table 1.** Selected baseline characteristics of the EPIC4ND case-cohort. Legend. This table presents selected baseline characteristics of the EPIC4ND case-cohort. Unless otherwise specified, all variables reflect the status at the time of recruitment. N=number, IQR=interquartile range, AAE=age at examination (recruitment), AAD=age at diagnosis, AAC=age at censoring, BMI=body mass index. Alcohol drinking pattern (lifestime):

| Disease | AD |  | Dementia |  | PD |  | ALS |  |
| --- | --- | --- | --- | --- | --- | --- | --- | --- |
| Case status | Cases | Non-cases | Cases | Non-cases | Cases | Non-cases | Cases | Non-cases |
| **N (% women)** | 818 (69) | 1818 (62) | 1190 (67) | 1785 (62) | 534 (48) | 4009 (65) | 199 (57) | 4585 (66) |
| **Median AAE (IQR)** | 59 (55-63) | 49 (43-56) | 59 (55-63) | 49 (43-55) | 60 (55-64) | 52 (46-59) | 58 (53-65) | 52 (46-59) |
| **Median AAD (IQR)** | 78 (74-81) |  | 78 (73-81) |  | 70 (64-75) |  | 73 (67-77) |  |
| **Median AAC (IQR)** |  | 71 (65-77) |  | 71 (65-77) |  | 69 (63-76) |  | 70 (64-77) |
| **Median follow-up time (IQR)** | 19 (16-22) | 22 (21-23) | 19 (16-21) | 22 (21-24) | 10 (6-14) | 16 (15-18) | 14 (9-18) | 18 (16-20) |
| **High/medium certainty, n (%)** | 801 (98) |  | 1117 (94) |  | 367 (69) |  | n.a. |  |
| **Education level, n (%)** |  |  |  |  |  |  |  |  |
| No education | 495 (61) | 639 (35) | 722 (61) | 616 (35) | 52 (10) | 500 (12) | 15 (8) | 692 (15) |
| Primary school | 229 (28) | 705 (39) | 330 (28) | 699 (39) | 187 (35) | 1505 (38) | 68 (34) | 1618 (35) |
| Technical/professional school | 25 (3) | 140 (8) | 38 (3) | 139 (8) | 116 (22) | 721 (18) | 47 (24) | 782 (17) |
| Secondary school | 32 (4) | 107 (6) | 38 (3) | 107 (6) | 70 (13) | 617 (15) | 20 (10) | 654 (14) |
| Longer education | 31 (4) | 214 (12) | 49 (4) | 213 (12) | 81 (15) | 566 (14) | 31 (16) | 683 (15) |
| Missing or not specified | 6 (1) | 13 (1) | 13 (1) | 11 (1) | 28 (5) | 100 (2) | 18 (9) | 156 (3) |
| **Smoking status, n (%)** |  |  |  |  |  |  |  |  |
| Never | 622 (76) | 1007 (55) | 865 (73) | 984 (55) | 267 (50) | 1901 (47) | 89 (45) | 2268 (49) |
| Former | 88 (11) | 324 (18) | 149 (13) | 320 (18) | 177 (33) | 1038 (26) | 69 (35) | 1161 (25) |
| Current | 108 (13) | 487 (27) | 175 (15) | 481 (27) | 78 (15) | 998 (25) | 33 (17) | 1084 (24) |
| Missing | 0 (0) | 0 (0) | 1 (0) | 0 (0) | 12 (2) | 72 (2) | 8 (4) | 72 (2) |
| **Coffee consumption (ml/d), n (%)** |  |  |  |  |  |  |  |  |
| 0 | 146 (18) | 202 (11) | 214 (18) | 196 (11) | 50 (9) | 297 (7) | 22 (11) | 360 (8) |
| 0.1-199.9 | 570 (70) | 1267 (70) | 827 (69) | 1243 (70) | 274 (51) | 2121 (53) | 77 (39) | 2470 (54) |
| 200-399.9 | 61 (7) | 265 (15) | 95 (8) | 262 (15) | 58 (11) | 548 (14) | 27 (14) | 596 (13) |
| 400-799.9 | 16 (2) | 43 (2) | 19 (2) | 43 (2) | 110 (21) | 720 (18) | 48 (24) | 792 (17) |
| >800 | 1 (0) | 4 (0) | 1 (0) | 4 (0) | 27 (5) | 220 (5) | 21 (11) | 253 (6) |
| Missing | 24 (3) | 37 (2) | 34 (3) | 37 (2) | 15 (3) | 103 (3) | 4 (2) | 114 (2) |
| **Alcohol drinking pattern (lifetime), n (%)** |  |  |  |  |  |  |  |  |
| Never | 249 (30) | 347 (19) | 343 (29) | 343 (19) | 31 (6) | 406 (10) | 12 (6) | 549 (12) |
| Former light | 112 (14) | 216 (12) | 162 (14) | 212 (12) | 55 (10) | 311 (8) | 15 (8) | 367 (8) |
| Former heavy | 9 (1) | 22 (1) | 18 (2) | 21 (1) | 3 (1) | 30 (1) | 2 (1) | 31 (1) |
| Light | 90 (11) | 193 (11) | 120 (10) | 188 (11) | 57 (11) | 424 (11) | 25 (13) | 541 (12) |
| Never heavy | 240 (29) | 702 (39) | 358 (30) | 690 (39) | 308 (58) | 1951 (49) | 105 (53) | 2171 (47) |
| Periodically heavy | 91 (11) | 270 (15) | 149 (13) | 263 (15) | 48 (9) | 461 (11) | 17 (9) | 496 (11) |
| Always heavy | 15 (2) | 58 (3) | 28 (2) | 58 (3) | 4 (1) | 70 (2) | 4 (2) | 72 (2) |
| Missing | 12 (1) | 10 (1) | 12 (1) | 10 (1) | 28 (5) | 356 (9) | 19 (10) | 358 (8) |
| **Physical activity, n (%)** |  |  |  |  |  |  |  |  |
| Inactive | 397 (49) | 675 (37) | 551 (46) | 659 (37) | 152 (28) | 1012 (25) | 51 (26) | 1248 (27) |
| Moderately inactive | 241 (29) | 595 (33) | 378 (32) | 582 (33) | 184 (34) | 1306 (33) | 63 (32) | 1513 (33) |
| Moderately active | 107 (13) | 325 (18) | 163 (14) | 321 (18) | 99 (19) | 803 (20) | 41 (21) | 884 (19) |
| Active | 73 (9) | 223 (12) | 98 (8) | 223 (12) | 81 (15) | 776 (19) | 37 (19) | 826 (18) |
| Missing | 0 (0) | 0 (0) | 0 (0) | 0 (0) | 18 (3) | 112 (3) | 7 (4) | 114 (2) |
| **BMI (kg/m2), n (%)** |  |  |  |  |  |  |  |  |
| <18.5 | 1 (0) | 1 (0) | 1 (0) | 1 (0) | 7 (1) | 22 (1) | 3 (2) | 27 (1) |
| 18.5-24.9 | 79 (10) | 411 (23) | 127 (11) | 405 (23) | 188 (35) | 1596 (40) | 96 (48) | 1765 (38) |
| 25-29.9 | 369 (45) | 872 (48) | 534 (45) | 857 (48) | 244 (46) | 1661 (41) | 70 (35) | 1910 (42) |
| >30 | 369 (45) | 534 (29) | 528 (44) | 522 (29) | 95 (18) | 730 (18) | 30 (15) | 883 (19) |
| **Diabetes, n (%)** |  |  |  |  |  |  |  |  |
| Yes | 89 (11) | 101 (6) | 153 (13) | 95 (5) | 27 (5) | 135 (3) | 6 (3) | 167 (4) |
| No | 729 (89) | 1714 (94) | 1037 (87) | 1687 (95) | 355 (66) | 3257 (81) | 122 (61) | 3796 (83) |
| Missing | 0 (0) | 3 (0) | 0 (0) | 3 (0) | 152 (28) | 617 (15) | 71 (36) | 622 (14) |
| **Hypertension, n (%)** |  |  |  |  |  |  |  |  |
| Yes | 260 (32) | 347 (19) | 386 (32) | 338 (19) | 145 (27) | 737 (18) | 39 (20) | 856 (19) |
| No | 557 (68) | 1470 (81) | 802 (67) | 1446 (81) | 226 (42) | 2228 (56) | 83 (42) | 2688 (59) |
| Missing | 1 (0) | 1 (0) | 2 (0) | 1 (0) | 163 (31) | 1044 (26) | 77 (39) | 1041 (23) |
| **Hyperlipidemia, n (%)** |  |  |  |  |  |  |  |  |
| Yes | 235 (29) | 382 (21) | 348 (29) | 371 (21) | 112 (21) | 694 (17) | 36 (18) | 798 (17) |
| No | 577 (71) | 1423 (78) | 835 (70) | 1402 (79) | 272 (51) | 2681 (67) | 95 (48) | 3145 (69) |
| Missing | 6 (1) | 13 (1) | 7 (1) | 12 (1) | 150 (28) | 634 (16) | 68 (34) | 642 (14) |
| **Cardiovascular problem, n (%)** |  |  |  |  |  |  |  |  |
| Yes | 268 (33) | 359 (20) | 397 (33) | 350 (20) | 159 (30) | 787 (20) | 42 (21) | 909 (20) |
| No | 548 (67) | 1455 (80) | 790 (66) | 1431 (80) | 341 (64) | 2730 (68) | 134 (67) | 3183 (69) |
| Missing | 2 (0) | 4 (0) | 3 (0) | 4 (0) | 34 (6) | 492 (12) | 23 (12) | 493 (11) |

However, data from these latter individuals can and have already been included in epidemiological, non-molecular studies investigating the entire EPIC cohort (see examples below).

The dementia/AD, PD, and ALS case ascertainments have been described previously^6–8,10,11^ (see **Supplementary Material** for details): Briefly, incident PD and dementia cases were first identified by record linkage with existing health databases and subsequently validated by re-review of medical records. Since the initial ascertainment, updates in selected centers yielded 435 additional incident dementia and 163 incident PD cases. For comparison purposes, 72 prevalent PD cases and 118 cases with Parkinsonism other than PD, which are not considered to be part of the main EPIC4ND dataset, have also been included in the molecular profiling (**Supplementary Table 5**). For ALS, cases were identified based on mortality records due to the fatal character of the disease and the short survival time after diagnosis.^6,11^ Since the original publications,^6,11^ the linkage to mortality records was updated centrally at IARC and/or locally for selected centers.

In addition to the EPIC4ND case-cohort, a French PD and ALS case-cohort (257 cases [238 PD, 19 ALS] and 281 non-cases) and a nested AD case-control dataset (n=352 incident cases, n=352 controls, with 142 additional longitudinal samples from cases) from Umeå, Sweden, are available as an extended molecular data resource within the EPIC4ND framework. Genomic and methylomic data have been generated for the French dataset and will be generated for the Swedish dataset using the same platforms and standardized data processing protocols as for the core EPIC4ND cohort. The generation of proteomic data in both datasets is planned as a next step.

### Omics data measurements

In the EPIC4ND case-cohort, proteomic data were generated from baseline plasma samples, while genome-wide DNA methylation and SNP genotyping data were generated from DNA extracted from buffy coats. The generation of small RNA sequencing data and targeted biomarker measurements from plasma samples is underway. In subsets of the nested PD and ALS case-control datasets, mass spectrometry has been performed on erythrocytes to also measure metals (PD and ALS)^12,13^ and on plasma to measure endogenous/biological metabolites and environmental chemicals (PD)^14,15^.

Specific to EPIC4ND, proteomic data were generated from plasma samples using the affinity-based SomaScan v4.1 assay (SomaLogic, Inc.) that measures ∼7,000 proteins. After quality control (QC), a total of 7,285 aptamers (6,381 proteins) across 1,859 ND cases and 4,417 non-cases are now available for statistical analysis. Methylomic data were generated from DNA extracted from buffy coats using the Infinium MethylationEPIC (v2) array (Illumina, Inc.) that contains ∼930k CpG sites. Following QC, 889,259 of these CpG sites are available for analysis across 1,659 ND cases and 2,586 non-cases. Genome-wide SNP genotyping data were generated using the Global Screening Array v1 (Illumina), which includes custom content for ∼40k mostly infrequent genetic variants located in genetic loci relevant for ND and movement disorders. Following QC and genotype imputation using established protocols,^16^ the final dataset comprises 7,381,754 SNPs in 1,637 ND cases and 2,598 non-cases available for statistical analysis. The “multi-*omics*” core dataset comprises 962 dementia (thereof 650 AD), 455 PD, and 170 ALS cases with data available on all three molecular domains. Due to differing numbers of EPIC centers per disease entity, the numbers of non-cases in the core dataset are 972 for the AD-specific EPIC4ND case-cohort (EPIC4AD), 2,226 for EPIC4PD, and 2,587 for EPIC4ALS (**Supplementary Tables 1-4**).

Furthermore, small RNA sequencing in plasma samples of 3,992 EPIC4ND participants (small RNA libraries prepared from the NextFlex Small-RNA-Seq kit (PerkinElmer, Waltham, Massachusetts), followed by sequencing on a HiSeq 4000 instrument (Illumina, San Diego, California) with 2×50 bp reads; data analysis will largely follow the protocol described in ref. ^17^) and targeted biomarker measurements in plasma samples of 1,700 EPIC4ND participants (measuring 120 biomarkers using the CNS NULISA assay, Alamar, Inc.) of the core dataset are currently being generated. In addition, in two overlapping nested case-control studies for PD and ALS (PD: n=362 cases, n=362 matched controls; ALS: n=107 cases, n=319 matched controls), data on metals in erythrocyte samples were generated using inductively coupled plasma mass spectrometry.^12,13^ Furthermore, for the nested case-control study in PD, metabolomics in plasma were generated using using both liquid chromatography-high resolution mass spectrometry (LC-HRMS) ^14,15^ and gas chromatography-high resolution mass spectrometry (GC-HRMS). LC-HRMS is effective for measuring endogenous/biological metabolites, while both LC-HRMS and GC-HRMS are capable of detecting internal markers for environmental chemicals. Most samples from these nested case-control studies also have corresponding genomic and methylomic data available, as described above. Finally, a large-scale whole exome/genome sequencing project, involving the sequencing of approximately 200,000 EPIC samples, is being planned as a future initiative. This project will also encompass EPIC4ND and the French dataset.

## Key findings

The EPIC4ND case-cohort comprises 1,899 incident ND cases (1,190 dementia thereof 818 AD, 534 PD, 199 ALS) and 4,447 non-cases across 12 centers from five European countries. The mean age at baseline was 54 (range 35-79) years for all EPIC4ND participants. The mean age at diagnosis for the incident cases was 77 (±7 standard deviation [SD]) years for dementia and 77±6 years for AD, 69±8 years for PD. Mean age at death for ALS (used as a proxy for age at diagnosis, see above) was 72±9 years. The mean time to diagnosis was 19±4 years for AD, 10±5 years for PD, and 13±6 years for ALS. The mean follow-up time for non-cases (defined as time until the last record of absence of the specific disease, usually at the time of last case ascertainment), was 22±4 years in EPIC4AD, 16±4 i years n EPIC4PD, and 18±5 years in EPIC4ALS.

In EPIC4AD, there were more women among cases than among non-cases. Future AD and other dementia cases were more likely to consume less coffee than average at recruitment, to be overweight, and have diabetes. Additionally, they tended to have lower educational attainment, be never smokers at recruitment, be physically inactive, and to be less likely to drink alcohol lightly compared to never drinkers but more likely to drink heavily. In EPIC4PD, cases comprised more men than non-cases. Future PD cases were less likely to smoke at recruitment and were more likely to be former light drinkers compared to never drinkers. Finally, in EPIC4ALS, there were more men among cases than among non-cases. Future ALS cases were less likely to be overweight (**Table 1, Figure 2, Supplementary Table 6**). These results are largely in line with previous EPIC cohort studies on dementia and AD^10^, PD (e.g., ref. ^18^) and ALS (e.g., ref. ^19^) using the entire EPIC cohort. Furthermore, the positive associations of AD risk with obesity and diabetes, and heavy alcohol consumption, and inverse association with education levels are in line with the major risk factors described in independent studies^20^ while the inverse association with smoking is unusual. In contrast, the inverse association of PD with smoking is well established based on numerous independent studies.^21^ This highlights that our smaller case-cohort design, optimized for molecular analyses, is not only representative of the larger study population but also accurately reflects many of the established risk factors in the source population.

**Figure 2.**
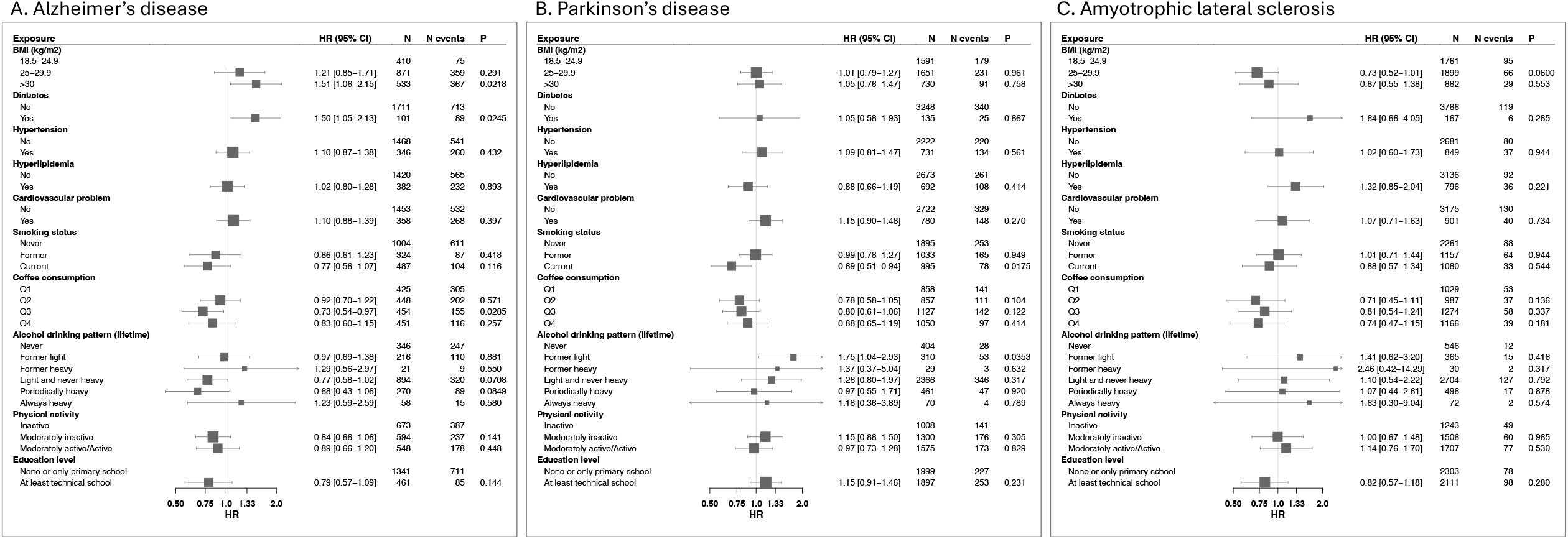
Forest plots of association analyses of baseline lifestyle and medical factors with instantaneous risk for Alzheimer’s disease, Parkinson’s disease and amyotrophic lateral sclerosis in the EPIC4ND case-cohort. Legend. These forest plots depict the association results of Prentice-weighted Cox proportional hazard regression analyses (stratified for age, sex, and center) on Alzheimer’s disease (A), Parkinson’s disease (B) and amyotrophic lateral sclerosis (C). HR=hazard ratio, CI=confidence interval, N=number, P=p-value, BMI=body mass index

The validity of our dataset is also substantiated by genome-wide association studies (GWAS) that we conducted in EPIC4ND using logistic regression analyses of AD, PD, and ALS (additive model, adjusted for the first four principal components, sex, and age at baseline). These analyses confirmed that the vast majority of EPIC4ND participants is of European descent (**Supplementary Figure 1**). These analyses revealed at least nominally significant effects of various well-established ND risk loci^22–25^, including *APOE* rs429358 (OR=4.02, p=4.73E-24) for AD, *GBA* rs35749011 (OR 2.27, p=1.00-02), *SNCA* rs356182 (OR 0.83, p=2.43-02) and *MAPT* rs62053943 (OR=0.79, p=4.28E-02) for PD, and *C9ORF72* rs2453555 (OR=1.37,p=1.13E-02) for ALS (**Supplementary Table 7, Supplementary Figure 2**). Several other known GWAS risk loci showed nominally significant effects (**Supplementary Table 7**). Furthermore, polygenic risk scores (PGS) for AD, PD, and ALS generated in the EPIC4ND case-cohort based on the established independent genome-wide significant (α=5×10-8) GWAS risk SNPs^22–25^ revealed strong associations with AD, PD, and ALS status. Specifically, we observed an OR of 2.04, 1.35, and 1.43 for AD, PD, and ALS, respectively, per one SD increase of the PGS (**Supplementary Table 8**). This supports the robustness of the case asertainment as well as the quality of the genetic data.

Furthermore, initial findings on plasma metabolites generated using mass spectrometry and on metals in erythrocytes have recently been published using overlapping, nested case-control studies for PD^12,14,15^ and ALS^13^.

## Strengths and limitations

This population-based case-cohort study is unique in the ND field for its study design with the availability of pre-disease blood samples, an extensive follow-up period of up to 30 years, and the new, in-depth measurements of multi-layered molecular data. Furthermore, EPIC4ND ensures high diagnostic accuracy by validating dementia, AD and PD cases through a re-review of medical records by expert neurologists following an initial case ascertainment via data linkage. This is in contrast to many other case-cohort studies and most mega-cohort studies with baseline biomaterial available, such as the UK Biobank, that rely solely on data linkage to study incident neurodegenerative diseases. Moreover, all study participants underwent extensive phenotyping and risk factor assessment, in particular diet, at baseline, yielding a comprehensive range of lifestyle and medical variables. In addition and despite being a multi center study, both the phenotypic characterization and the processes for retrieving and storing blood samples have been highly standardized and harmonized, minimizing between-center heterogeneity. Notwithstanding, our study has several weaknesses: First, although we aimed for the highest possible quality in ascertainment of AD, PD, and ALS cases within our study, the accuracy of the diagnosis ultimately lies in the responsibility of the diagnosing or treating physician, and there may be some variability across centers. We cannot rule some degree of case misclassification; underascertainment, in particular, is more likely to exist for AD. Second, while we aimed to obtain high-quality diagnostic information standardized evaluations of specific disease phenotypes (e.g., initial symptoms) and of disease progression (e.g., severity or therapy response) are largely missing. Lastly, EPIC is based in Europe and as demonstrated above the vast majority of participants are of European descent. As a result, although the ascertainment of a pan-European cohort is a strength in terms of generalizability to European populations, the generalizability of findings to other ethnic groups and populations is limited, highlighting the urgent need for future studies including more diverse populations.

## Conclusions

EPIC4ND is a unique resource for identifying novel molecular biomarkers for a future onset of AD, PD, and ALS and for investigating biomarkers and pathways shared across ND and other diseases. Additionally, it enables the investigation of the interplay and interactions among multiple omics domains. Its validity is supported by associations with established lifestyle and genetic risk factors. Similar studies are urgently needed to address the critical need for identifying individuals at high risk of ND, facilitating the development of targeted preventive and early therapeutic interventions.

## Data Availability

All data produced in the present study are available upon reasonable request to the authors

## Acknowledgements

The authors thank all participants of the project and the countless scientists who have contributed to the EPIC cohort over the past 30 years. We sincerely thank Bertrand Hemon for his assistance with data management, Tanja Wesse for technical support, and Prof. Paolo Vineis, Prof. Beate Ritz, Prof. Martin Wolkewitz, Prof. Klaus Berger and Prof. Valentina Gallo for helpful discussions.

## Data access

Data can be accessed by external researchers provided the proposed project is approved by the EPIC4ND working group and the EPIC steering committee. Potential collaborators can contact the EPIC4ND working group chair, Christina Lill, EPIC administrator Sherry Morris, and EPIC PI Elio Riboli.

## Statements and Declarations

## Funding

The updates of the case ascertainments, generation and processing of the molecular data were funded by the Michael J Fox Foundation (#008994 to C.M.L. and E.R.), the Cure Alzheimer’s Fund (to C.M.L. and L.B.), the ‘CReATe-Clinical Research in ALS and Related Disorders for Therapeutic Development’ Consortium (to C.M.L. and L.B.), a grant from the EU Joint Programme – Neurodegenerative Disease Research (JPND2021-650-289, coordinator: C.M.L.), and the Deutsche Forschungsgemeinschaft (DFG; LI 2654/3-1, to C.M.L.). The case ascertainment for the dementia cases in Granada was funded by the Instituto de Investigación Biosanitaria ibs.GRANADA (INTRAIBS-2020-06). C.M. Lill was supported by the Heisenberg program of the DFG (DFG; LI 2654/4-1). See **Supplementary Material** for additional funding information and a disclaimer.

## Competing interests

H.Z. has served at scientific advisory boards and/or as a consultant for Abbvie, Acumen, Alector, Alzinova, ALZpath, Amylyx, Annexon, Apellis, Artery Therapeutics, AZTherapies, Cognito Therapeutics, CogRx, Denali, Eisai, Enigma, LabCorp, Merry Life, Nervgen, Novo Nordisk, Optoceutics, Passage Bio, Pinteon Therapeutics, Prothena, Quanterix, Red Abbey Labs, reMYND, Roche, Samumed, Siemens Healthineers, Triplet Therapeutics, and Wave, has given lectures sponsored by Alzecure, BioArctic, Biogen, Cellectricon, Fujirebio, Lilly, Novo Nordisk, Roche, and WebMD, and is a co-founder of Brain Biomarker Solutions in Gothenburg AB (BBS), which is a part of the GU Ventures Incubator Program. None of the other authors reports any relevant conflict of interests.

## Author contributions

C.M.L., M.G., and E.R., conceived the study with help from S.G., L.M., L.B., and P.F.; C.M.L., J.H., K.S.-B., V.V., J.M.H., A.B., V.D., O.M., M.G., D.P., M.T., J.S., M.G., S.M. C.-Y., S.M., N. C.-C., S.E., S.S., A.J., T.Y.N.T., R.K., G.S., R.T., N.W., A.L.B., H.Z., A.F., A.E., L.B., R.V., L.M., G.M., C.S., S.P., V.K., P.F., M.G., and E.R. carried out data collection; C.M.L., J.H., O.O., K. S.-B., V.V., Y.Z., F.A., L.D., O.R., A.E., L.B., P.F., E.R. and M.G. carried out analysis and/or interpretation of data. C.M.L. and J.H. drafted the manuscript; all authors critically revised the manuscript for intellectual content. All authors read and approved the final manuscript of the paper.

## Ethics approval

The study was approved by the ethical committee of the International Agency for Research on Cancer (IARC) and by the ethical review boards of participating study centers.

## Consent to participate

All participants provided written informed consent.

## Consent to publish

Not applicable.

## References

1. Riboli E, Kaaks R. The EPIC Project: rationale and study design. European Prospective Investigation into Cancer and Nutrition. Int J Epidemiol. 1997;26 Suppl 1:S6–14. doi:10.1093/ije/26.suppl_1.s6

2. Riboli E, Hunt KJ, Slimani N, et al. European Prospective Investigation into Cancer and Nutrition (EPIC): study populations and data collection. Public Health Nutr. 2002;5(6B):1113–1124. doi:10.1079/PHN2002394

3. InterAct Consortium, Langenberg C, Sharp S, et al. Design and cohort description of the InterAct Project: an examination of the interaction of genetic and lifestyle factors on the incidence of type 2 diabetes in the EPIC Study. Diabetologia. 2011;54(9):2272–2282. doi:10.1007/s00125-011-2182-9

4. Danesh J, Saracci R, Berglund G, et al. EPIC-Heart: the cardiovascular component of a prospective study of nutritional, lifestyle and biological factors in 520,000 middle-aged participants from 10 European countries. Eur J Epidemiol. 2007;22(2):129–141. doi:10.1007/s10654-006-9096-8

5. Lopes EW, Chan SSM, Song M, et al. Lifestyle factors for the prevention of inflammatory bowel disease. Gut. Published online December 6, 2022. doi:10.1136/gutjnl-2022-328174

6. Gallo V, Bueno-De-Mesquita HB, Vermeulen R, et al. Smoking and risk for amyotrophic lateral sclerosis: analysis of the EPIC cohort. Ann Neurol. 2009;65(4):378–385. doi:10.1002/ana.21653

7. Gallo V, Brayne C, Forsgren L, et al. Parkinson’s Disease Case Ascertainment in the EPIC Cohort: The NeuroEPIC4PD Study. Neurodegener Dis. 2015;15(6):331–338. doi:10.1159/000381857

8. Andreu-Reinón ME, Gavrila D, Chirlaque MD, et al. Ascertainment of Dementia Cases in the Spanish European Prospective Investigation into Cancer and Nutrition-Murcia Cohort. Neuroepidemiology. 2019;52(1-2):63–73. doi:10.1159/000493209

9. Rafii MS. Preclinical Alzheimer’s disease therapeutics. J Alzheimers Dis. 2014;42 Suppl 4:S545–9. doi:10.3233/JAD-141482

10. Andreu-Reinón ME, Huerta JM, Gavrila D, et al. Incidence of Dementia and Associated Factors in the EPIC-Spain Dementia Cohort. J Alzheimers Dis. 2020;78(2):543–555. doi:10.3233/JAD-200774

11. Gallo V, Vanacore N, Bueno-de-Mesquita HB, et al. Physical activity and risk of Amyotrophic Lateral Sclerosis in a prospective cohort study. Eur J Epidemiol. 2016;31(3):255–266. doi:10.1007/s10654-016-0119-9

12. Zhao Y, Ray A, Broberg K, et al. Prediagnostic Blood Metal Levels and the Risk of Parkinson’s Disease: A Large European Prospective Cohort. Mov Disord. 2023;38(12):2302–2307. doi:10.1002/mds.29602

13. Peters S, Broberg K, Gallo V, et al. Blood Metal Levels and Amyotrophic Lateral Sclerosis Risk: A Prospective Cohort. Ann Neurol. 2021;89(1):125–133. doi:10.1002/ana.25932

14. Zhao Y, Lai Y, Darweesh SKL, et al. Gut Microbial Metabolites and Future Risk of Parkinson’s Disease: A Metabolome-Wide Association Study. Mov Disord. Published online November 12, 2024. doi:10.1002/mds.30054

15. Zhao Y, Lai Y, Konijnenberg H, et al. Association of Coffee Consumption and Prediagnostic Caffeine Metabolites With Incident Parkinson Disease in a Population-Based Cohort. Neurology. 2024;102(8):e209201. doi:10.1212/WNL.0000000000209201

16. Hong S, Prokopenko D, Dobricic V, et al. Genome-wide association study of Alzheimer’s disease CSF biomarkers in the EMIF-AD Multimodal Biomarker Discovery dataset. Transl Psychiatry. 2020;10(1):403. doi:10.1038/s41398-020-01074-z

17. Dobricic V, Schilling M, Schulz J, et al. Differential microRNA expression analyses across two brain regions in Alzheimer’s disease. Transl Psychiatry. 2022;12(1):352. doi:10.1038/s41398-022-02108-4

18. Gallo V, Vineis P, Cancellieri M, et al. Exploring causality of the association between smoking and Parkinson’s disease. Int J Epidemiol. 2019;48(3):912–925. doi:10.1093/ije/dyy230

19. Gallo V, Wark PA, Jenab M, et al. Prediagnostic body fat and risk of death from amyotrophic lateral sclerosis: the EPIC cohort. Neurology. 2013;80(9):829–838. doi:10.1212/WNL.0b013e3182840689

20. Livingston G, Huntley J, Sommerlad A, et al. Dementia prevention, intervention, and care: 2020 report of the Lancet Commission. Lancet. 2020;396(10248):413–446. doi:10.1016/S0140-6736(20)30367-6

21. Noyce AJ, Bestwick JP, Silveira-Moriyama L, et al. Meta-analysis of early nonmotor features and risk factors for Parkinson disease. Ann Neurol. 2012;72(6):893–901. doi:10.1002/ana.23687

22. Nalls MA, Blauwendraat C, Vallerga CL, et al. Identification of novel risk loci, causal insights, and heritable risk for Parkinson’s disease: a meta-analysis of genome-wide association studies. Lancet Neurol. 2019;18(12):1091–1102. doi:10.1016/S1474-4422(19)30320-5

23. Lambert JC, Ibrahim-Verbaas CA, Harold D, et al. Meta-analysis of 74,046 individuals identifies 11 new susceptibility loci for Alzheimer’s disease. Nat Genet. 2013;45(12):1452–1458. doi:10.1038/ng.2802

24. Bellenguez C, Küçükali F, Jansen IE, et al. New insights into the genetic etiology of Alzheimer’s disease and related dementias. Nat Genet. 2022;54(4):412–436. doi:10.1038/s41588-022-01024-z

25. van Rheenen W, van der Spek RAA, Bakker MK, et al. Common and rare variant association analyses in amyotrophic lateral sclerosis identify 15 risk loci with distinct genetic architectures and neuron-specific biology. Nat Genet. 2021;53(12):1636–1648. doi:10.1038/s41588-021-00973-1

